# Synchronous remote-based interventions for suicidal behaviour prevention: a systematic review and meta-analyses of clinical trials

**DOI:** 10.1101/2024.05.21.24306868

**Authors:** Laura Comendador, María P. Jiménez-Villamizar, Josep-Maria Losilla, Juan P. Sanabria-Mazo, Corel Mateo-Canedo, Antoni Sanz, Ana-Isabel Cebrià, Diego Palao

**Affiliations:** Department of Psychiatry and Forensic Medicine, Faculty of Medicine, Universitat Autònoma de Barcelona. 08193 Cerdanyola del Vallès, Spain; Department of Mental Health, Parc Taulí University Hospital. Institut d’Investigació i Innovació Parc Taulí (I3PT- CERCA). Universitat Autònoma de Barcelona. 08208 Sabadell, Spain; Department of Basic, Developmental, and Educational Psychology, Faculty of Psychology, Universitat Autònoma de Barcelona. 08193 Cerdanyola del Vallès, Spain; Department of Psychobiology and Methodology of Health Sciences, Faculty of Psychology, Universitat Autònoma de Barcelona. 08193 Cerdanyola del Vallès, Spain; Teaching, Research & Innovation Unit, Parc Sanitari Sant Joan de Déu. 08830 Sant Boi de Llobregat, Spain; Centre for Biomedical Research in Epidemiology and Public Health (CIBERESP). 28029 Madrid, Spain; Stress and Health Research Group (GIES). Universitat Autònoma de Barcelona. 08193 Cerdanyola del Vallès, Spain; Centro de Investigación Biomédica en Red de Salud Mental (CIBERSAM), Instituto de Salud Carlos III. 28029 Madrid, Spain; Department of Clinical and Health Psychology, Faculty of Psychology, Universitat Autònoma de Barcelona. 08193 Cerdanyola del Vallès, Spain

**Keywords:** Suicide, Telemedicine, Preventive Medicine

## Abstract

**Objectives:** Suicide is a leading cause of preventable death worldwide. Evidence supports the impact of providing active contact for individuals who have attempted suicide. The current systematic review and meta-analyses aim to investigate the effects of suicide prevention strategies implemented through remote and synchronous technology-based interventions.

**Design:** Systematic review, narrative synthesis, and meta-analysis.

**Data sources:** Electronic databases (PubMed, PsycInfo, Scopus, and Web of Science) and grey literature sources (ClinicalTrials.gov and Google Scholar) were searched until December 2024. **Eligibility criteria** Eligible articles assessed suicide prevention interventions for participants over 12 years with prior suicidal behaviour. Eligible study design included randomised control trials and non-randomised clinical trials published in English or Spanish.

**Data extraction and synthesis:** Screening, selection process, data extraction, and risk of bias assessment were performed independently by two reviewers. Data on suicide-related factors and adherence to treatment were extracted. Meta-analyses were conducted to determine effect sizes (Hedges’ *g*) for suicidal ideation, risk ratios (RR) for suicide attempts, and Peto odds ratios (OR) for suicide. Heterogeneity was assessed using the Cochrane’s Q test, tau^2^ statistic, and *I*^2^ value. Publication bias was investigated employing funnel plots and Egger’s test.

**Results:** A total of 28 studies, comprising 10,015 participants in the intervention group and 10,726 in the comparison group, were included in the systematic review and meta-analyses. Synchronous remote-based interventions were effective in preventing repeated suicide attempts at 1 month (RR 0.73, 95% CI 0.62 to 0.85, *I*^2^=0.0%, Q=0.70, tau^2^=0.00), 6 months (RR 0.56, 95% CI 0.34 to 0.95, *I*^2^=85.4%, Q=54.92, tau^2^=0.36), and 12 months (RR 0.68, 95% CI 0.49 to 0.96, *I*^2^=87.6%, Q=72.63, tau^2^=0.27). Additionally, these interventions were associated with a reduction in suicide-related deaths at 18 months (Peto OR 0.18, 95% CI 0.08 to 0.44, *I*^2^=0.0%, Q=0.03, tau^2^=0.00). Effects on suicidal ideation were not statistically significant at any time-point (Hedges’ *g* -0.07 to -0.28, *I*^2^=0.0 to 69.3%, Q=1.16 to 7.38, tau^2^=0.00 to 0.14).

**Conclusions:** Synchronous remote-based interventions demonstrate a potential benefit in preventing suicide attempts and deaths by suicide, and may serve as an adjunct to usual treatment; however, the effect on suicidal ideation appears limited. The observed heterogeneity warrants caution when interpreting these findings. Future research should prioritise methodological enhancements to improve the quality and consistency of evidence, as well as investigate the mediating processes underlying their effectiveness in reducing suicidal behaviour.

**PROSPERO registration number** CRD42021275044.

**STRENGTHS AND LIMITATIONS OF THIS STUDY:**

- The protocol was registered in PROSPERO and published in a peer-reviewed journal, ensuring methodological transparency.
- Data extraction, synthesis, and quality assessment are reported in accordance with the Preferred Reporting Items for Systematic Reviews and Meta-Analyses (PRISMA-2020) guidelines.
- The systematic review and meta-analyses used a comprehensive search strategy to avoid missing published articles.
- Heterogeneity in the methods makes it difficult to draw definitive conclusions about the effectiveness of interventions.

## INTRODUCTION

The World Health Organisation (WHO) estimates that over 703,000 people die annually from suicide worldwide [1, 2]. Suicide occurs across the lifespan; although, in 2019, it was the fourth leading cause of death among individuals aged 15 – 29 years worldwide [2, 3]. For each death by suicide, there are twenty suicide attempts [4], which is one of the major disease burden causes in the world [5, 6, 7]. Moreover, a history of suicide attempts is considered a robust predictor of completed suicide, supporting prevention efforts during the acute period following an episode of suicidal behaviour [8–12].

Increasing evidence suggests that post-discharge follow-up contacts can be an effective suicide prevention strategy [13–19]. The eHealth-based treatments could be classified into two categories: (1) real-time synchronous therapy – such as telephone contact or telepsychiatry via videoconferencing – (2) or digital self-help programmes without human guidance, often asynchronous [20, 21]. Synchronous communication technologies, including telemedicine and real-time interactions, have the potential to provide immediate and flexible access to support during critical periods. As opposed to asynchronous methods, these approaches facilitate direct and immediate engagement between patients and healthcare professionals, potentially enhancing the effectiveness of suicide prevention efforts. Accordingly, most post-discharge suicide prevention programmes reinforce early contact with mental health professionals. An essential component of the effectiveness of suicide prevention interventions is treatment adherence [22]. The evaluation of therapeutic adherence in clinical care is critically important since poor adherence results in adverse outcomes, such as relapses and hospitalisations, among people with mental health disorders worldwide [22, 23, 24]. Brief contact interventions have three rationales: firstly, they allow for repeated suicide risk assessments; secondly, they reduce feelings of social isolation and provide emotional support; and thirdly, they improve treatment adherence and reduce dropout from mental health care [25–28].

In recent decades, numerous studies have reported promising results from the implementation of follow-up contact interventions for at-risk individuals as a suicide prevention strategy [29, 30]. Although previous systematic reviews have addressed this topic [31, 32, 33], evidence on the specific impact of synchronous technologies remains limited. In a scoping review, Shin *et al.* [34] identified several trends in information and communication technology (ICT)-based interventions for suicide prevention applied in clinical settings, such as computerised interventions or applications, highlighting the need for future research to strengthen the evidence to improve implementation. This review included 66 ICT-based interventions, targeted at post-discharge follow-up, screening, and safety planning, for any spectrum of suicide-related thoughts and behaviours.

Along these lines, Luxton *et al.* [35] suggest that repeated follow-up contacts may help reduce suicidal behaviour, reporting in their systematic review four studies that showed mixed results with trends towards a preventive effect and two studies that showed no preventive effect. A more recent systematic review and meta-analysis [36] examined 16 studies evaluating the effectiveness of telehealth interventions in suicide prevention, reporting that telepsychiatry is effective in reducing suicide and reattempt rates, with high acceptance and retention rates. Nevertheless, this review did not specifically evaluate suicidal ideation, and moreover, the authors emphasise the need for further research to fully elucidate the potential of telepsychiatry in suicide prevention.

These interventions’ distinctive and rapidly evolving nature necessitates a thorough exploration of their implications. Our research intends to expand on previous evidence by updating it with a larger number of studies, a comprehensive analysis of the components of suicidal behaviour, and a more detailed examination of the temporal dynamics of the effects. The current systematic review and meta-analyses aim to explore the evidence on remote suicide prevention strategies implemented through synchronous technology-based interventions (i.e., via remote, interactive, real-time communication), considering the effects, barriers, and facilitators to the implementation.

## METHODS

### Protocol and registration

The protocol of the systematic review and meta-analyses was registered in the Prospective International Registry of Systematic Reviews (PROSPERO), with the identification number CRD42021275044. The review was reported according to the Preferred Reporting Items for Systematic Reviews and Meta-Analyses (PRISMA) guidelines [37] (see Supplemental File 1: Table S1). A brief mention of the amendments made to the original protocol article [38] can be found in the Supplemental File 2.

### Eligibility criteria

The main inclusion and exclusion criteria are described below. Full details on the eligibility criteria can be found in the protocol article [38].

#### Participants

Adolescents over 12 years of age and adults with reported suicidal ideation or prior suicide attempts. Individuals with non-suicidal self-injury were excluded.

#### Interventions

Secondary suicide prevention interventions delivered via synchronous distance communication technologies– i.e., telephone contact, 24-hour telephone hotlines, videoconferencing therapy, immediate text messaging, etc–. Interventions based on asynchronous telecommunication devices were excluded.

#### Comparisons

The eligible control groups include treatment as usual (e.g., visits to mental health services or pharmacotherapy), enhanced usual care, no treatment, historical control, or geographical control. Non-reporting of a control group was a reason for exclusion. Studies were excluded if both, the intervention and control groups, received synchronous and remote interventions.

#### Outcomes

Suicidal behaviours– i.e., suicidal ideation, suicide attempts, and suicide deaths– were included. Suicidal ideation refers to thoughts about the possibility of ending one’s life [39], assessed using validated instruments such as the Columbia Suicide Severity Rating Scale (CSSRS). A suicide attempt is defined as a self-inflicted, potentially injurious act with a non-fatal outcome and evidence of intent to die [3], measured by the number of reported attempts within a defined follow-up period. Finally, suicide death involves a self-inflicted behaviour performed with the deliberate intention of resulting in death [40], and may be quantified by the number of persons who die by suicide. Outcomes could be assessed at any point following the intervention, with no restriction on the length of follow-up, provided the assessment uses a quantitative approach. Adherence to treatment was also extracted as a secondary outcome.

#### Study design

Randomised controlled trials (RCTs) and non-randomised clinical trials (nRCTs) were eligible for inclusion. The term nRCTs refers to studies with a defined intervention and comparison group, in the absence of random allocation. Observational studies uncontrolled and non-experimental were excluded (i.e., single-arm studies, interrupted time series, case reports, descriptive non- comparative studies) [41, 42].

### Data collection and analysis

#### Information sources

Bibliographic searches were conducted in PubMed, PsycInfo, Scopus, and Web of Science for studies published before December 2024. ClinicalTrials.gov and Google Scholar were searched for grey literature and unpublished records. Reference lists of previous systematic reviews on the subject and references cited in eligible articles were consulted for data collection.

#### Search strategy

Details of the search process applied to databases, registers, and websites can be found in Supplemental File 3. The search was unrestricted by year of publication, limited to English or Spanish, and developed adhering to the Peer Review of Electronic Search Strategies (PRESS) [43].

#### Study selection

The results of the literature search were imported into Mendeley (version 1.19.8), and Rayyan Systems Inc [44]. Duplicate articles were removed automatically by Rayyan Systems Inc. and manually by the first reviewer (LC). After, two authors (LC and MPJ) blind-screened all articles based on titles, abstracts, and keywords. Then, two reviewers (LC and MPJ) independently evaluated the full-text articles according to the eligibility criteria. Discrepancies were resolved through discussion with a third author (AS) until reaching consensus.

#### Data collection

Two authors (LC and MPJ) performed the data extraction independently, applying an *ad hoc* extraction form in Microsoft Excel (version 16.65).

#### Data items

Data were extracted on participants and intervention characteristics, along with outcomes related to suicidal behaviour for both intervention and comparison groups– for all measures and time points–. In addition, therapeutic adherence– specifically, the moderating effect of adherence on the effectiveness of the intervention, the description of measures to maximise adherence to treatment, and predictor variables of adherence– were examined.

### Risk of bias

Two reviewers (LC and MPJ) performed the risk of bias (RoB) assessment independently, using the Revised Cochrane risk-of-bias tool for randomised trials (RoB 2) [45] and Risk-of-bias In Non-randomised Studies of Interventions (ROBINS-I) [46] for the outcome of suicidal behaviours. Disagreements were resolved by consensus with a third blinded reviewer (AS).

### Effect measures

Systematically examined all types of effect measures used in the selected trials (e.g., risk ratio, odds ratio, mean difference, frequencies) for the effects of each intervention group on suicidal ideation, repeated suicide attempts, and death by suicide.

### Synthesis methods

Firstly, a systematic review was employed to collect and synthesise the empirical evidence that met the eligibility criteria. Secondly, meta-analyses were conducted using a random-effects model. Due to the wide variety of study types included in the systematic review, a random-effects model was selected *a priori* for all analyses, and heterogeneity statistics were reported using this model, in accordance with the recommendations from the Cochrane Handbook for Systematic Reviews of Interventions (version 6.3) [47]. To ensure that the combined numerical data from the different studies is comparable and can be adequately synthesised, analyses by sorting studies with the same evaluation measures time frame were performed for each outcome, and a narrative synthesis approach was adopted when a quantitative combination of the studies was not feasible.

Three types of meta-analyses were performed. (1) A random-effects model with Hedges’ *g* [48] and 95% confidence intervals (CI) were calculated for suicidal ideation. Effect sizes are considered small (*g*≥0.2), medium (*g*≥0.5), or large (*g*≥0.8) [48]. (2) A random-effects model with the effect estimates weighted by the inverse of the variance was calculated for suicide attempts. As proposed by Daly [49] and Zhang & Yu [50], the risk ratio (RR) and 95% CI were analysed. Finally, (3) a random-effects model with the effect estimates weighted by the Peto *et al.* method [51] was calculated for suicide deaths since the observed frequencies are very low. The Cochrane’s Q test and tau^2^ statistic were employed to identify heterogeneity among studies. The proportion of variability due to heterogeneity was indicated by *I*^2^ value. Heterogeneity was considered moderate if *I*^2^ was greater than 30%. All analyses were performed using the Meta-Essentials tool for Microsoft Excel (version 16.65) [52], applying the Knapp-Hartung adjustment by default in fitting random effects models. The trials with multiple assessment measurement periods were included as separate studies in the appropriate meta-analyses [47].

### Certainty of evidence

The certainty of evidence was assessed using the Grading of Recommendations Assessment, Development, and Evaluation (https://gdt.gradepro.org/) [53]. The GRADE guidelines establish standardised and transparent criteria for grading the certainty of evidence [54]. In classifying the level of evidence quality, GRADE defines four categories: high, moderate, low, and very low [55]. The main domains used are risk of bias, inconsistency, indirect evidence, imprecision, and publication bias [55]. Two reviewers (LC and MPJ) independently evaluated the certainty of the evidence of the included studies; any discrepancies were resolved through consultation with a third reviewer (AS). The assessment was carried out for each outcome of suicidal behaviours. Summary tables of the findings were presented to synthesise the process.

### Patient and public involvement

Not involved in the review protocol, design, reporting, or dissemination plans.

## RESULTS

### Study selection

The initial search retrieved 4930 studies. After removing duplicates, 2802 articles were screened for title, abstract, and keywords. Of these, 131 studies were selected for full-text review. Finally, 28 publications were included in the systematic review and meta-analyses [56–83]. The article selection process was described in a PRISMA flowchart [84] (Figure 1). Studies that might appear to meet the inclusion criteria but were excluded and the reasons for exclusion are presented in Supplemental File 4: Table S2. Inter-rater agreement was calculated for initial screening using Delta (*δ*=0.91, 95% CI 0.88 to 0.94) [85], and for eligibility and extraction process using Cohen’s Kappa (*κ*=0.82, 95% CI 0.62 to 1.0; *κ*=0.98, 95% CI 0.97 to 0.99, respectively) [86].

**Figure 1.**
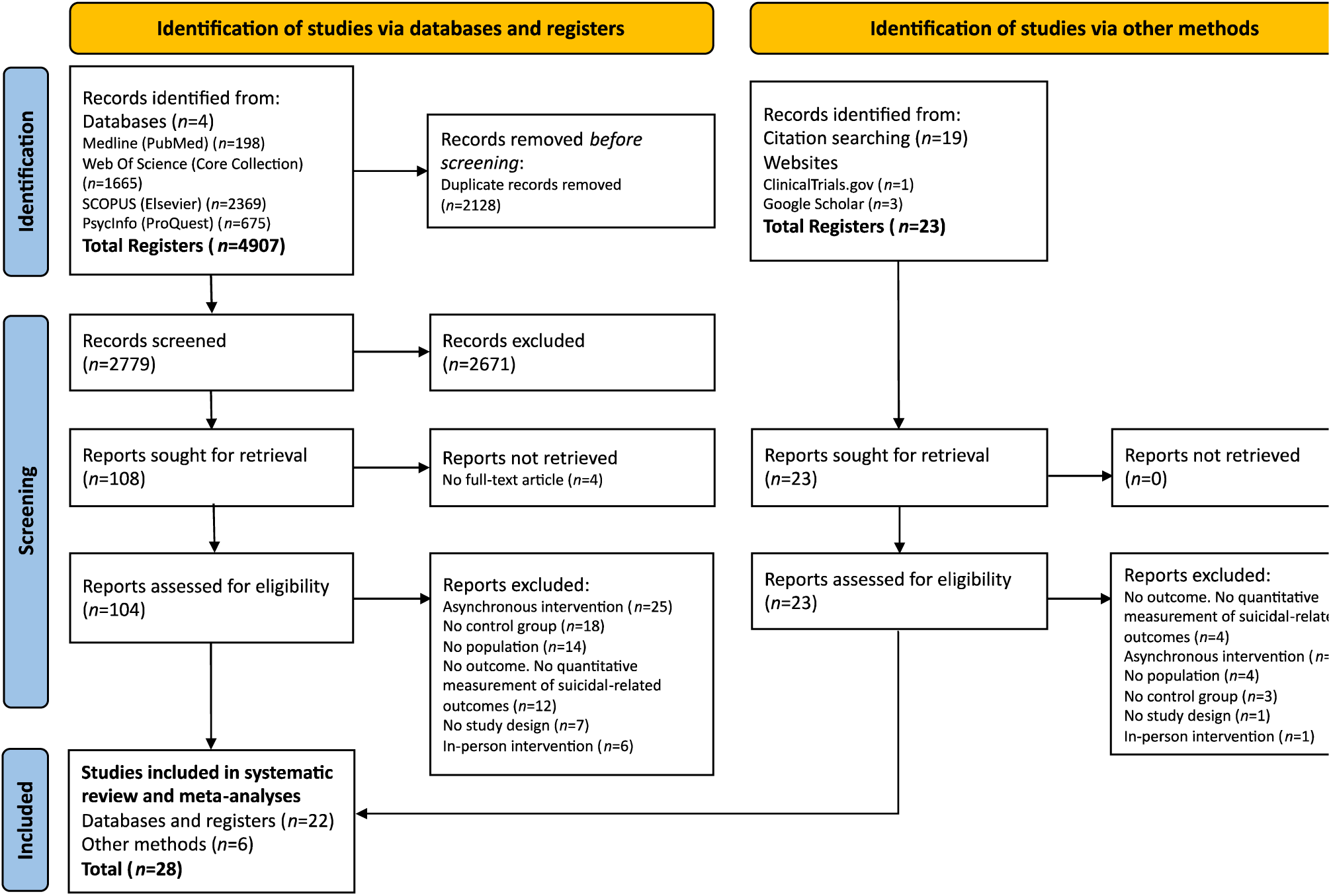
PRISMA flowchart for systematic review and meta-analyses including database searches and other sources.

### Study characteristics

A descriptive summary of the characteristics of the included studies is presented in Table 1 and Supplemental File 5: Table S3. The current systematic review included 28 studies, of which 19 were RCT (67.9%) [56–58, 61–63, 66, 69, 71, 72, 74–77, 79–83]. The sample size ranges from 10 to 3068 in the intervention group (IG) and from 10 to 3694 in the control group (CG). The mean age ranged from 23 to 45 years in the IG and from 23 to 47 years in the CG. The proportion of women in most of the studies was above 50% in both IG and CG, except in three studies [63, 79, 82]. The eligible articles were from 12 countries: France [64, 65, 72, 74, 78, 89, 81], Spain [59, 60, 67, 68, 70], United States of America [63, 73, 79, 82], Iran [69, 71, 75, 76], Brazil, India, Sri Lanka [57, 66], French Polynesia [56], Canada [58], Sweden [61], Australia [62], United Kingdom [77], and China [83].

**Table 1:** Overview of participants, interventions, comparisons, outcomes, and study design (PICOS) for studies evaluating suicide ideation, attempted suicide, and suicide death.

| Study (authors, date of publication, country) | Study design | Setting, sample size (N) | Details of participants | Intervention (n) and comparison (n) | Follow-up | Outcome measure (Instrument) |
| --- | --- | --- | --- | --- | --- | --- |
| Amadéo *et al.*, 2015 [56]. French Polynesia | RCT | Psychiatric ED<br>N=190 | Patients with suicidal behaviour; mean age IG 33, CG 31.48; 64.2% women | BIC (n=90) vs. TAU (n=100). 1h psycho-educational session combined with telephone contacts at 1, 2, 4, 7, 11 weeks, 4, 6, 12, and 18 months by psychologists and psychiatrists. | 18 months | Suicide attempt (EPSIS) [87], Suicide death (Forensic records of suicide deaths) |
| Bertolote *et al.*, 2010 [57]. Brazil, India, Sri Lanka, Iran, China | RCT | ED<br>N=1867 | Patients with suicide attempt; mean age IG 26.3, CG 25.5; 58.2% women | BIC (n=922) vs. TAU (n=945). 1h brief educational intervention combined with telephone contacts or visits at 1, 2, 4, 7, 11 weeks, 4, 6, 12, 18 months by medical doctor, psychiatric nurse, psychologist, or social worker. | 18 months | Suicide attempt (EPSIS) [87] |
| Burback *et al.*, 2024 [58]. Canada | RCT | Mental health services. N=42 | Patients with suicidal ideation in the past week; age IG 36.0, CG 36.0; 79.0% women | Web-based therapist-delivered EMDR (n=20) vs. TAU (n=22). 12 sessions of web-based EMDR (90 min.) delivered twice weekly via videoconferencing by psychiatrists. | 4 months | Suicidal ideation (BSS) [88], Suicide attempt (CSSRS) [89], Suicide death (CSSRS) [89] |
| Cebrià *et al.*, 2013 [59]. Spain | nRCT; case-control | 2 Hospital ED<br>N=514 | Patients discharged from ED after a suicide attempt; age IG 41.92, CG 40.73; 66.5% women | Telephone follow-up (n=296) vs. TAU (n=218). Telephone contacts (5-10 min.) at 1 week, 1, 3, 6, 9, and 12 months by a psychiatric nurse. | 12 months | Suicide attempt (Interview + medical record) |
| Cebrià *et al.*, 2015 [60]. Spain | nRCT; retrospective cohort | 2 Hospital ED<br>N=485 | Patients discharged from ED after a suicide attempt; age IG 38.78, CG 41.92; 64.5% women | Telephone follow-up (n=287) vs. TAU (n=198). Telephone contacts (5-10 min.) at 1 week, 1, 3, 6, 9, and 12 months by a psychiatric nurse. | 60 months | Suicide attempt, Suicide death (Interview + Medical records) |
| Cedereke *et al.*, 2002 [61]. Sweden | RCT | Hospital, Emergency Unit<br>N=216 | Patients treated after a suicide attempt; mean age IG 40, CG 42; 66% women | Telephone follow-up (n=107) vs. TAU (n=109). Telephone contacts (20-25 min.) at 4 and 8 months after a suicide attempt, by a psychiatric nurse or social counsellor. | 12 months | Suicidal ideation (SSI) [90], Suicide attempt (Semi-structured interview) |
| Christensen *et al.*, 2013 [62]. Australia | RCT | Australia's Lifeline. N=155 | Callers to a national 24h-helpline service with moderate-to-high psychological distress; mean age 41.49; 81.9% women | Web-based CBT (n=38) vs. Web-based CBT + telephone follow-up (n=45) vs. telephone follow-up (n=37) vs. TAU (n=35). CBT delivered across five modules. Telephone contacts (10 min.), weekly, by a Lifeline telephone counsellor. | 12 months | Suicidal ideation (GHQ-28) [91] |
| Comtois *et al.*, 2019 [63]. USA | RCT | 3 military installations<br>N=657 | Soldiers and marines identified as being at risk of suicide; mean age IG 25.6, CG 24.8; 17.9% women | Caring contacts (n=329) vs. TAU (n=328). Text messages delivered on day 1, at week 1, at months 1, 2, 3, 4, 6, 8, 10, and 12, and on participants' birthdays, by mental health clinicians. Text message replies from participants were monitored 24 hours a day. Replies indicating suicide risk or distress were handled by a few text messages exchanged or a single telephone call. | 12 months | Suicidal ideation (SSI) [90], Suicide attempt (Interview + SASI-Count) [92], Suicide death (Interview) |
| Demesmaeker *et al.*, 2024 [64]. France | nRCT; prospective cohort | General medical or psychiatric services<br>N=733 | Patients discharged from the hospital after a suicide attempt (non-first-time attempters); mean age IG 40.2, CG 39.4; 61.5% women | Telephone contact (n=236) vs. Telephone contact and postcards (n=411) vs. Postcards (n=34) vs. TAU (n=52). Telephone contact between the 10<sup>th</sup> and 21<sup>st</sup> days after discharge for suicide attempts by psychologists. Personalised postcards are sent at months 2–5 to those unavailable for a phone call or facing a suicidal crisis. | 6 months | Suicide attempt (Standardised telephone interview) |
| Exbrayat *et al.*, 2017 [65]. France | nRCT; non-randomised controlled | Emergency Psychiatry Dept.<br>N=827 | Patients admitted to the ED for suicide attempt; mean age IG 40.2, CG 39.7; 69.5% women | Telephone follow-up (n=436) vs. TAU (n=387). Telephone contacts at 8, 30, 60 days after suicide attempt by nurses. | 2 months | Suicide attempt (medical records) |
| Fleischmann *et al.*, 2008 [66]. Brazil, India, Sri Lanka, Iran, China | RCT | ED<br>N=1867 | Patients admitted to the ED for suicide attempt; mean age IG 23, CG 23; 58.2% women | BIC (n=922) vs. TAU (n=945). 1h brief educational intervention combined with telephone contacts (5 min.) or visits at 1, 2, 4, 7, 11 weeks, 4, 6, 12, and 18 months by a medical doctor, psychiatric nurse, psychologist, or social worker. | 18 months | Suicide death (EPSIS) [87] |
| Gabilondo *et al.*, 2020 [67]. Spain | nRCT; non-randomised controlled | 3 General Hospitals<br>N=586 | Patients discharged from a General Hospital after a suicide attempt; mean age IG 41.2, CG 45.2; 58.5% women | Telephone follow-up (n=123) vs. TAU (n=463). 6 months telephone contacts (10-15 min.) after 1, 2 weeks, 1, 3, 6 months by general nurses. | 12 months | Suicide attempt (medical records) |
| Goñi-Sarriés *et al.*, 2022 [68]. Spain | nRCT; prospective cohort | 2 Psychiatric Units<br>N=191 | Patients admitted to the ED after attempted suicide; mean age IG 44.96, CG 46.91; 52.9% women | Telephone follow-up (n=101) vs. TAU (n=90). Telephone contacts (15-20 min.) the day after discharge, at 15 days, and at 2, 4, 8, and 12 months by a specialist nurse. | 12 months | Suicide attempts, Suicide death (Interview) |
| Hassanzadeh *et al.*, 2010 [69]. Iran | RCT | 5 General Hospitals ED<br>N=632 | Patients with suicide attempt; mean age IG 24 (SD=8.3), mean age CG 25; 62% women | BIC (n=319) vs. TAU (n=310). 1-hour psycho-educational session combined with telephone contacts at 1, 2, 4, 7, 11 weeks, 4, and 6 months by psychologists, nurses, consultants, and psychiatric residents. | 6 months | Suicide attempts, Suicide death (SUPRE-MISS questionnaire) [93] |
| López-Goñi & Goñi-Sarriés 2021 [70]. Spain | nRCT; prospective cohort | Psychiatric ED<br>N=410 | Patients discharged from the ED for a suicide attempt; mean age IG 41.7, CG 44.6; 63.7% women | Telephone follow-up (*n*=203) vs. TAU (*n*=207). Telephone follow-up at 15 days, 2, 4, 8, and 12 months by mental health nurses. | 12 months | Suicide attempt, Suicide death (medical records) |
| Malakouti *et al.*, 2022 [71]. Iran | RCT | Hospital ED<br>N=305 | Patients admitted to the ED after attempted suicide; 66.9% women | Telephone contact programme (*n*=153) vs. TAU (*n*=152). Telephone contact calls (15-20 min.) at 1, 2, and 4 weeks, and monthly for up to 12 months by clinical psychologists with a master's degree. | 12 months | Suicide attempts, Suicide death (SUPRE-MISS questionnaire) [93] |
| Messiah *et al.*, 2019 [72]. France | RCT | 23 Hospital ED and psychiatric crisis centres<br>N=1040 | Patients discharged from the ED for a suicide attempt; 65.9% women | AlgoS: Telephone follow-up for multi-attempters (*n*=317) or crisis card for first attempters (*n*=203) vs. TAU (*n*=520). Telephone follow-up between the 10<sup>th</sup> and 21<sup>st</sup> by a psychologist. The patient could not be contacted after 3 attempts, on 3 different days, at 3 different times of day: postcards were sent at 2, 3, 4, and 5 months. | 14 months | Suicide attempt (Semi-structured interview) |
| Miller *et al.*, 2017 [73]. USA | nRCT; non-randomised controlled | 8 Hospital ED<br>N=1376 | Patients with suicide attempt or ideation presented to hospital ED; mean age IG 36, CG 37; 56% women | Universal screening and ED-SAFE intervention (*n*=502) vs. Universal screening (*n*=377) vs. TAU (*n*=497). Risk screening by the ED physician, discharge resource, and 7 post-ED telephone calls (10-20 min.). | 12 months | Suicide attempt, Suicide death (CSSRS) [89] |
| Mouaffak *et al.*, 2015 [74]. France | RCT | Psychiatric ED<br>N=320 | Patients admitted to the ED for suicide attempt; mean age IG 39, CG 38.6; 70% women | Postal cards, 24-hour telephone and telephone follow-up (*n*=160) vs. TAU (*n*=160). Postal cards, few days, 1, 6, and 11 months providing 24-hour crisis telephone consultation with a psychiatrist, and telephone calls at 2 weeks, 1- and 3 months PD. | 12 months | Suicide attempt (SIS) [94], Suicide death (Semi-structured interview) |
| Mousavi *et al.*, 2014 [75]. Iran | RCT | Hospital ED<br>N=139 | Patients admitted to the ED for suicide attempt in 2010-2011; 63.3% women | Telephone follow-up (*n*=69) vs. TAU (*n*=70). Telephone contacts (30 min.) at 2, 4 weeks, 2, 3, 4, 5, and 6 months by psychiatry. | 6 months | Suicide attempt (*ad hoc* questionnaire) |
| Mousavi *et al.*, 2016 [76]. Iran | RCT | Hospital ED<br>N=55 | Patients admitted to ED for suicide attempt; mean age IG 27.07, CG 29.69; 87.3% women | Telephone follow-up (*n*=29) vs. face-to-face (*n*=26). Telephone contacts (20 min.) at 2, 4 weeks, 2, 3, 4, 5, 6, and 8 months by an assistant of psychiatry. | 8 months | Suicide attempt (Interview) |
| O'Connor *et al.*, 2022 [77]. UK | RCT | 4 general hospitals<br>N=120 | Patients with a history of a suicide attempt; mean age IG 36.1, CG 37.0 (*SD*=14.8); 62.5% women | SAFETEL intervention (*n*=80) vs. TAU (*n*=40). Safety planning intervention session and 5 optional telephone calls at 72h of patient discharge from the hospital and weekly thereafter depending on participants' availability by clinical psychologists. | 6 months | Suicide Ideation, Suicide attempt (CSSRS) [89] |
| Plancke *et al.*, 2020 [78]. France | nRCT; retrospective cohort | Hospital ED<br>N=6762 | Patients discharged from the hospital after a suicide attempt; mean age IG 37.9, mean age CG 39.4; 61.4% women | Telephone follow-up (*n*=3068) vs. TAU (*n*=3694). Telephone contacts at 10, 21 days, and 6 months by psychiatrists. The patient could not be reached by telephone: post-cards were sent at 2, 3, 4, and 5 months. | 6 months | Suicide attempt (Interview) |
| Riblet *et al.*, 2022 [79]. USA | RCT | Veterans Medical Center<br>N=20 | Patients at risk for suicide; mean age IG 54.9, CG 48.1; 35% women | BIC (*n*=10) vs. TAU (*n*=10). 6 contacts by phone or video over 3 months, educational materials mailed out to the patient, and 1-hour educational intervention on suicide prevention either over the phone or video by a clinical psychologist. | 3 months | Suicide Ideation (BSS) [88] + Suicide attempt (CSSRS) [89] |
| Vaiva *et al.*, 2006 [80]. France | RCT | 13 ED<br>N=605 | Patients discharged from an ED after attempted suicide by deliberate self-poisoning; mean age IG 37, CG 35; 72.9% women | Telephone contact 1 month (*n*=147) or 3 months (*n*=146) vs. TAU (*n*=312). Telephone contact (20-25 min.) at 1 or 3 months after discharge from an ED for attempted suicide by psychiatrists. | 13 months | Suicide attempts, Suicide death (medical records) |
| Vaiva *et al.*, 2018 [81]. France | RCT | 23 ED and psychiatry crisis centers<br>N=987 | Patients discharged from the ED for a suicide attempt; mean age IG 38.4 (*SD*=13.4), mean age CG 38.1 (*SD*=13.1); 63.4% women | AlgoS algorithm (*n*=493) vs. TAU (*n*=494). Telephone contacts between the 10th and 21st day after a suicide attempt by a psychologist. The patient could not be contacted after 3 attempts, on 3 different days, at 3 different times of day, or decline follow-up care from other services, or identified being under stress/experiencing a suicidal crisis: programmed mailing post-cards at months 2, 3, 4, and 5. | 13 months | Suicide attempts, Suicide death (Interview) |
| Ward-Ciesielski *et al.*, 2017 [82]. USA | RCT | University Outpatient Clinic. N=93 | Participants with suicidal ideation; mean age IG 38.6 (*SD*=15.0), mean age CG 41.8 (*SD*=15.3); 37% women | DBT-BSI and telephone follow-up (*n*=46) vs. RT control condition (*n*=47). Single-session DBT-BSI, presenting pre-selected skills (mindfulness, opposite-to-emotion action, distraction, and changing your body chemistry) and 3 months telephone contact (30-45 min.) at 1, 4, and 12 weeks. | 3 months | Suicidal ideation (SSI) [90], Suicide attempts (Interview) |
| Wei *et al.*, 2013 [83]. China | RCT | 4 General Hospitals ED<br>N=239 | Patients admitted to the ED after attempted suicide; mean age IG 34.06 (*SD*=15.84), mean age CG 32.12 (*SD*=13.87); 77.1% women | Cognitive Therapy group (*n*=82) vs. Telephone intervention group (*n*=80), (3) vs. TAU (*n*=77). 12 weekly phone calls (20-40 min.) within a 3-month period by professors. | 12 months | Suicide Ideation (SSI) [90], Suicide attempt (Structured Interview) |

As for outcome variables, 8 studies reported outcomes relating to suicidal ideation [58, 61–63, 77, 79, 82, 83], 26 studies reported the proportion of participants who reattempted suicide [56–61, 63–65, 67–83], and 13 reported the proportion of suicide deaths [56, 58, 60, 63, 66, 68–71, 73, 74, 80, 81]. There was high heterogeneity in the diagnostic methods used to identify suicide behaviours: European Parasuicide Study Interview Schedule (EPSIS), Scale for Suicide Ideation (SSI), General Health Questionnaire (GHQ-28), Columbia Suicide Severity Rating Scale (CSSRS), Beck Scale for Suicidal Ideation (BSS), Suicide Attempt Self-Injury Count (SASI- Count), Beck’s Suicide Intent Scale (SIS), Multisite Intervention Study on Suicidal Behaviours (SUPRE-MISS) questionnaire, forensic records of suicide deaths, data from electronic medical records, etc. The most frequently reported psychiatric diagnoses were affective disorders (*n*=13; 46.4%) [56, 58, 61, 64, 65, 68, 70, 72, 73, 74, 77, 79, 81], followed by anxiety disorders (*n*=10; 35.7%) [56, 58, 61, 64, 65, 72, 73, 77, 79, 81], personality disorders (*n*=5; 17.9%) [56, 58, 65, 68, 74], substance use disorders (*n*=5; 17.9%) [56, 58, 64, 72, 77], trauma-related disorders (*n*=5; 17.9%) [58, 64, 74, 79, 81], psychotic disorders (*n*=4; 14.3%) [56, 65, 68, 73], and eating disorders (*n*=3; 10.7%) [65, 72, 81] according to the Diagnostic and Statistical Manual of Mental Disorders (DSM-III-R, DSM-IV-TR) or the International Classification of Diseases (ICD-10).

The interventions were categorised as (a) telephone follow-up intervention (*n*=16; 57.1%) [59–61, 64, 65, 67, 68, 70–72, 75, 76, 78, 80, 81, 83], (b) Brief Intervention and Contact (BIC; *n*=5; 17.9%) [56, 57, 66, 69, 79], (c) web-based therapist-delivered Eye Movement Desensitization and Reprocessing (EMDR; *n*=1; 3.6%) [58], (d) 24-hour call-in services (*n*=1; 3.6%) [62], (e) instant text messages (*n*=1; 3.6%) [63], (f) Emergency Department Safety Assessment and Follow-up Evaluation (ED-SAFE intervention; *n*=1; 3.6%) [73], (g) postal cards, 24-hour telephone, and telephone follow-up (*n*=1; 3.6%) [74], (h) Safety Planning and telephone follow-up intervention (SAFETEL intervention; *n*=1; 3.6%) [77], and (i) Dialectical Behaviour Therapy Skills-Based Intervention (DBT-BSI) and telephone follow-up (*n*=1; 3.6%) [82]. On the other hand, the control group in most studies was TAU (*n*=26; 92.9%) [56–75, 77–81, 83], except for two studies, which was a face-to-face follow-up [76] and a relaxation training condition [82].

In relation to setting, the included studies were conducted in emergency departments (*n*=19; 67.9%) [56, 57, 59–61, 65, 66, 69–76, 78, 80, 81, 83], mental health services (*n*=4; 14.3%) [58, 64, 68, 82], general hospitals (*n*=2; 7.1%) [67, 77], military installations (*n*=2; 7.1%) [63, 79], or 24-hour call-in services (*n*=1; 3.6%) [62]. Interventions were performed by nurses (*n*=7; 25%) [59–61, 65, 67, 68, 70], psychologists (*n*=7; 25%) [63, 64, 71, 72, 77, 79, 81], psychiatrists (*n*=6; 21.4%) [58, 74–76, 78, 80], physicians (*n*=1; 3.6%) [73], and by a combination of health professionals– i.e., medical doctor, psychiatric nurse, psychologist, or social worker (*n*=4; 14.3%) [56, 57, 66, 69]– although, few interventions were undertaken by non-health professionals (*n*=2; 7.1%) [62, 83] or not reported (*n*=1; 3.6%) [82]. The onset of the intervention– i.e., initial contact– could be categorised in a time interval of days (*n*=5; 17.9%) [58, 63, 64, 74, 77], weeks (*n*=19; 67.9%) [56, 57, 59, 60, 62, 65–72, 75, 76, 78, 81–83], months (*n*=2; 7.1%) [61, 80], or not reported (*n*=2; 7.1%) [73, 79]. The total number of contacts ranged from one [64, 72, 80, 81] to 14 [71], with a minimum duration of five minutes [59, 60, 66] and a maximum of 90 minutes [58]. In addition, the follow-up period ranged from two months [65] to five years [60], with 12 months being the most prevalent period range (*n*=10; 35.7%) [59, 61–63, 67, 68, 70, 71, 73, 74].

### Risk of bias in studies

The results of the RoB 2 [45] and ROBINS-I [46] evaluations are presented in Figure 2. Inter-rater agreement was calculated using Cohen’s Kappa (*κ*=0.94, 95% CI 0.87 to 1.0) [86]. The overall RoB ratings were classified as low, moderate, or high. A total of 8 (28.6%) studies met the criteria to be classified as low risk of bias; in 13 (46.4%) studies the risk of bias was moderate; while in 7 (25%) studies the risk of bias was high for the outcome of suicidal behaviours.

**Figure 2.**
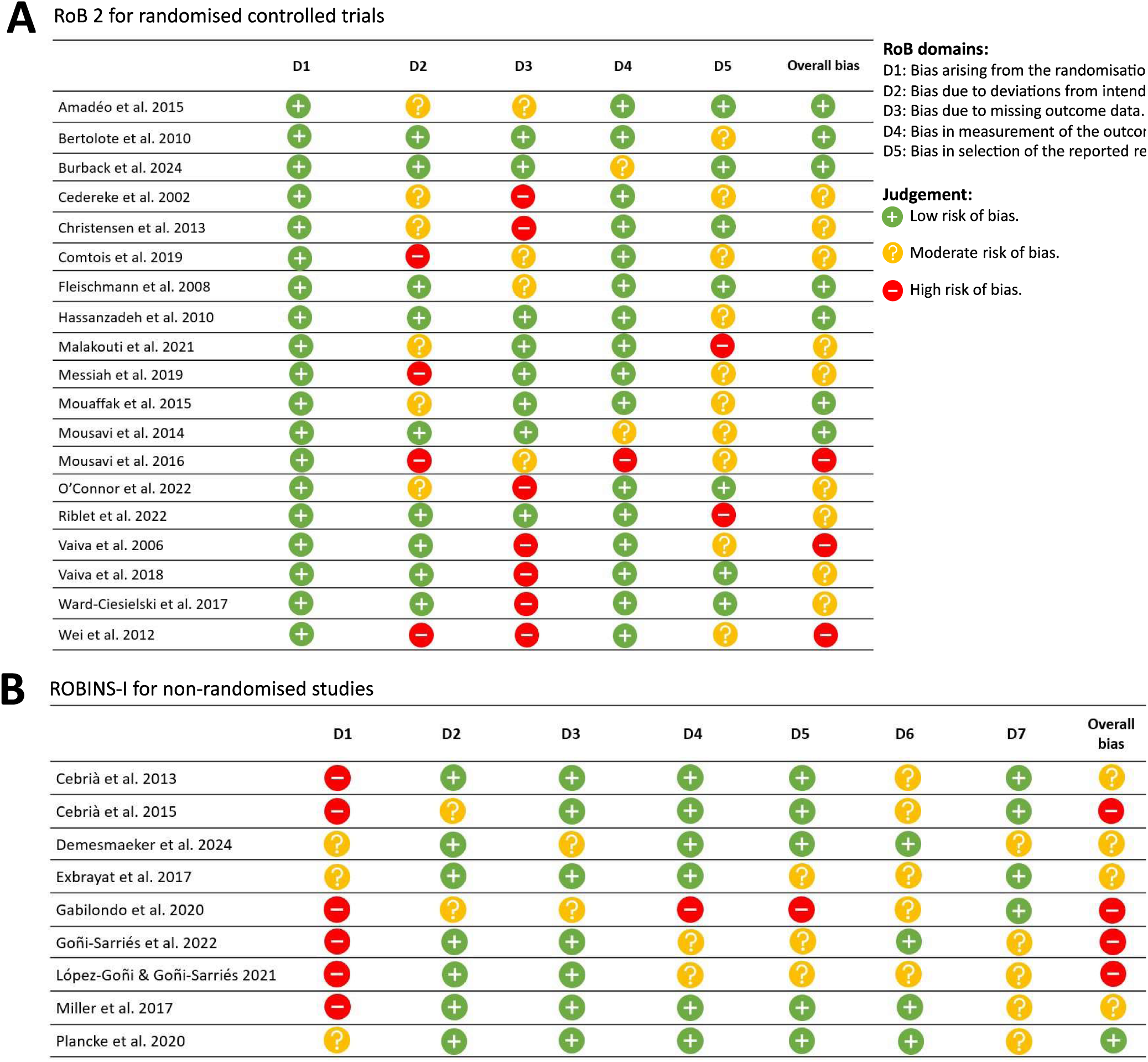
RoB assessment. (**A**) RoB 2 evaluation; (**B**) ROBINS-I evaluation.

### Results of individual studies

#### Suicidal ideation

The results of the meta-analyses of the effects of synchronous remote-based intervention on suicidal ideation are shown in Figure 3. Among the 28 trials, four studies reported the effects of the intervention on suicidal ideation in short-term (≤ 3 months) [58, 62, 79, 82] and four studies in medium-term (6–12 months) [61–63, 77]. There was a very small size, non-statistically significant effect of IG vs. CG on suicidal ideation at short-term (Hedges’ *g* -0.17, 95% CI -0.62 to 0.28 at 1 month; Hedges’ g -0.07, 95% CI -0.34 to 0.20 at 3 months). Also, there was a small size, non-statistically significant effect at medium-term (Hedges’ *g* -0.28, 95% CI -0.89 to 0.34 at 6 months; Hedges’ *g* -0.27, 95% CI -0.56 to 0.03 at 12 months). Heterogeneity was substantial for nearly all the measurement moments (*I*^2^=59.4% at 1 month, *I*^2^=69.3% at 6 months, *I*^2^=51.9% at 12 months) except for the measurement at 3 months (*I*^2^=0.0%).

**Figure 3.**
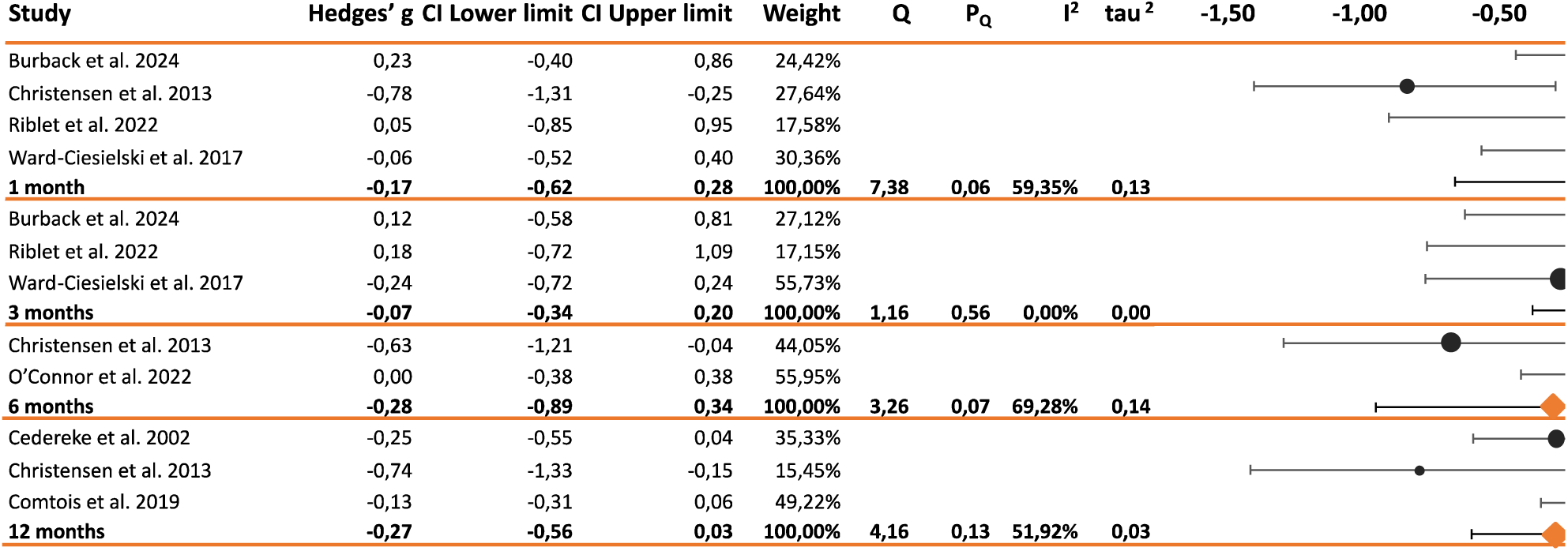
Forest plot comparing the effects of synchronous – remote interventions on suicidal intention.

#### Suicide attempts

The results of the meta-analyses of the effects of synchronous remote-based intervention on suicide reattempts are shown in Figure 4. Among the 28 trials, seven studies reported the effects of the intervention on suicide attempts in short-term (≤ 3 months) [58, 65, 75, 76, 79, 82, 83], 17 studies in medium-term (6–12 months) [59, 63, 64, 67–78, 81, 83], and six studies in long-term (>12 months) [56, 57, 61, 72, 80, 81]. There was a statistically significant effect (RR 0.73, 95% CI 0.62 to 0.85) on suicide attempts at 1 month favourable to the intervention group, with a 27% reduction of the risk of making at least one suicide attempt, relative to control group. There was a statistically significant effect (RR 0.56, 95% CI 0.34 to 0.95) on suicide attempts at 6 months, with a 44% lower risk of making at least one suicide attempt in the intervention group compared to the control group, despite considerable heterogeneity (*I*^2^=85.4%). In addition, a statistically significant effect (RR 0.68, 95% CI 0.49 to 0.96) on suicide attempts at 12 months was observed, with a 32% lower risk of making at least one suicide attempt in the intervention group compared to the control group, although there was considerable heterogeneity (*I*^2^=87.6%). For the remaining measurement points, the heterogeneity was non-obvious (*I*^2^=0.0% at 1 month, 3 months, 13 months, and 18 months).

**Figure 4.**
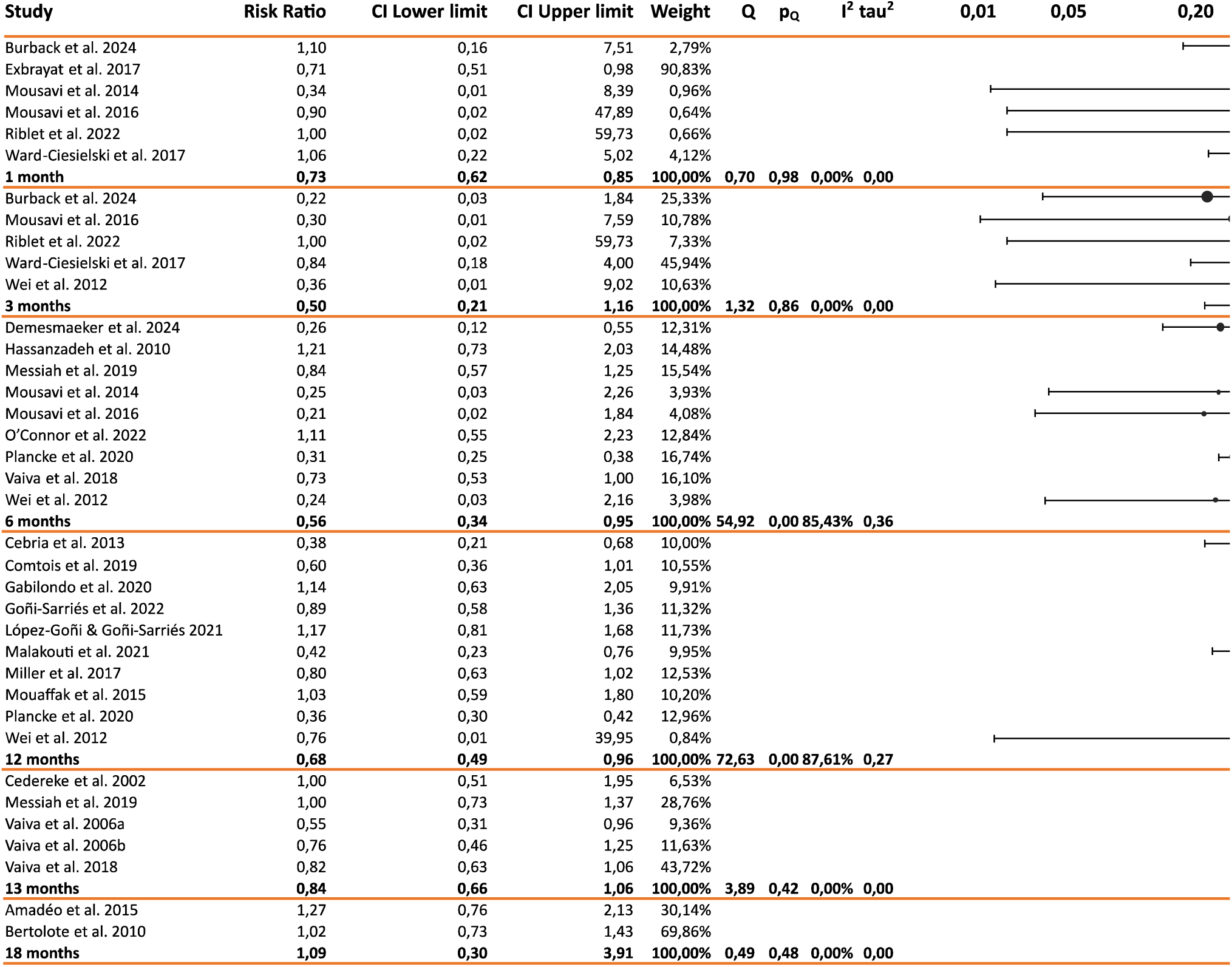
Forest plot comparing the effects of synchronous – remote interventions on suicide attempts.

#### Suicide deaths

The results of the meta-analyses of the effects of synchronous remote-based intervention on suicide deaths are shown in Figure 5. Among the 28 trials, seven studies reported the effects of the intervention on suicide deaths in medium-term (6–12 months) [63, 68–70, 73, 74, 81], and four studies in long-term (>12 months) [56, 66, 80, 81]. There was a statistically significant effect on suicide deaths at 18 months (Peto OR 0.18, 95% CI 0.08 to 0.44), with the likelihood of dying by suicide being 0.18 times lower in the intervention group compared to the control group. The heterogeneity was low for all measurement points (*I*^2^=17.9% at 6 months, *I*^2^=0.0% at 12 months, 13 months, and 18 months).

**Figure 5.**
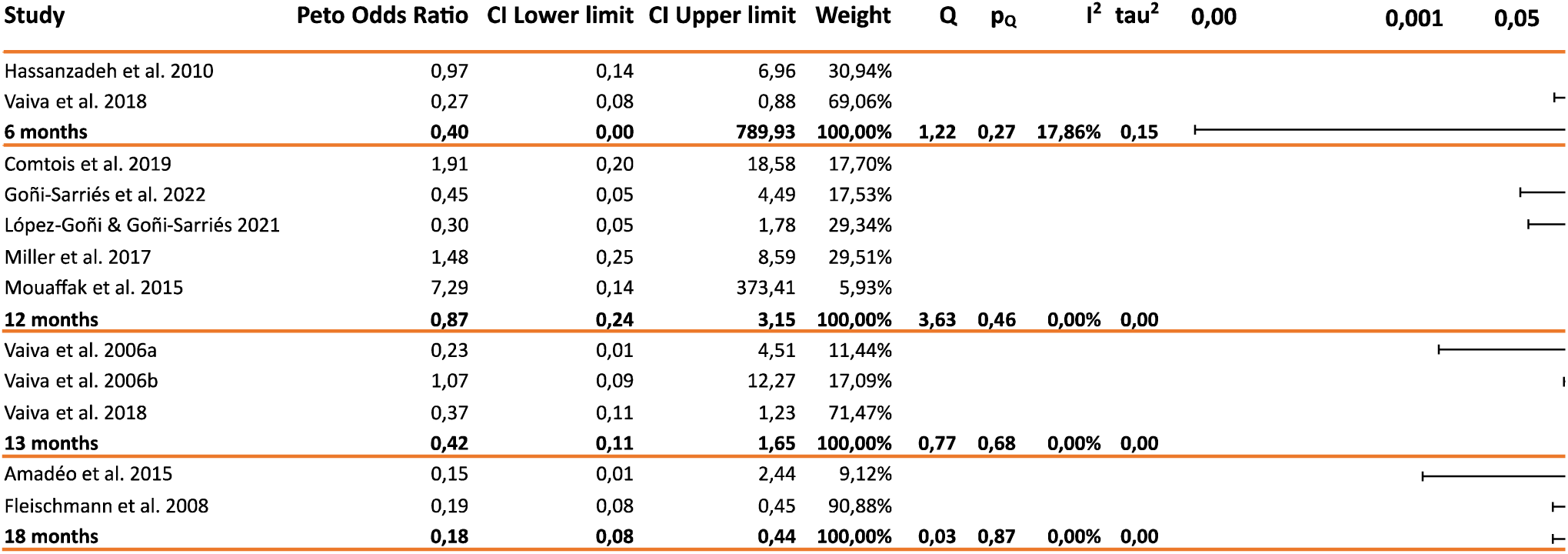
Forest plot comparing the effects of synchronous – remote interventions on suicide deaths.

In order to simplify interpretation, the results can be grouped into three timepoints: short- (≤ 3 months), medium- (6–12 months), and long-term (>12 months) (Table 2).

**Table 2:**
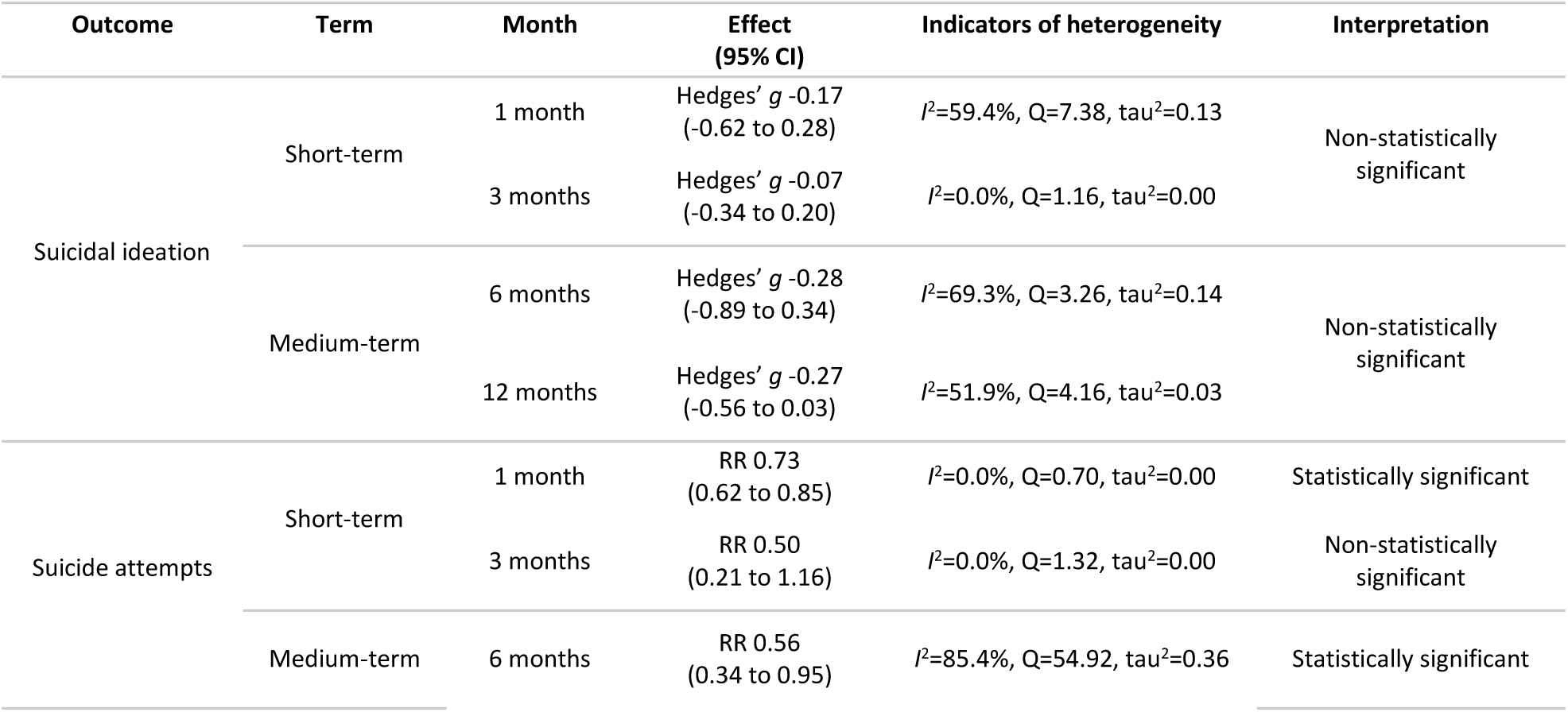
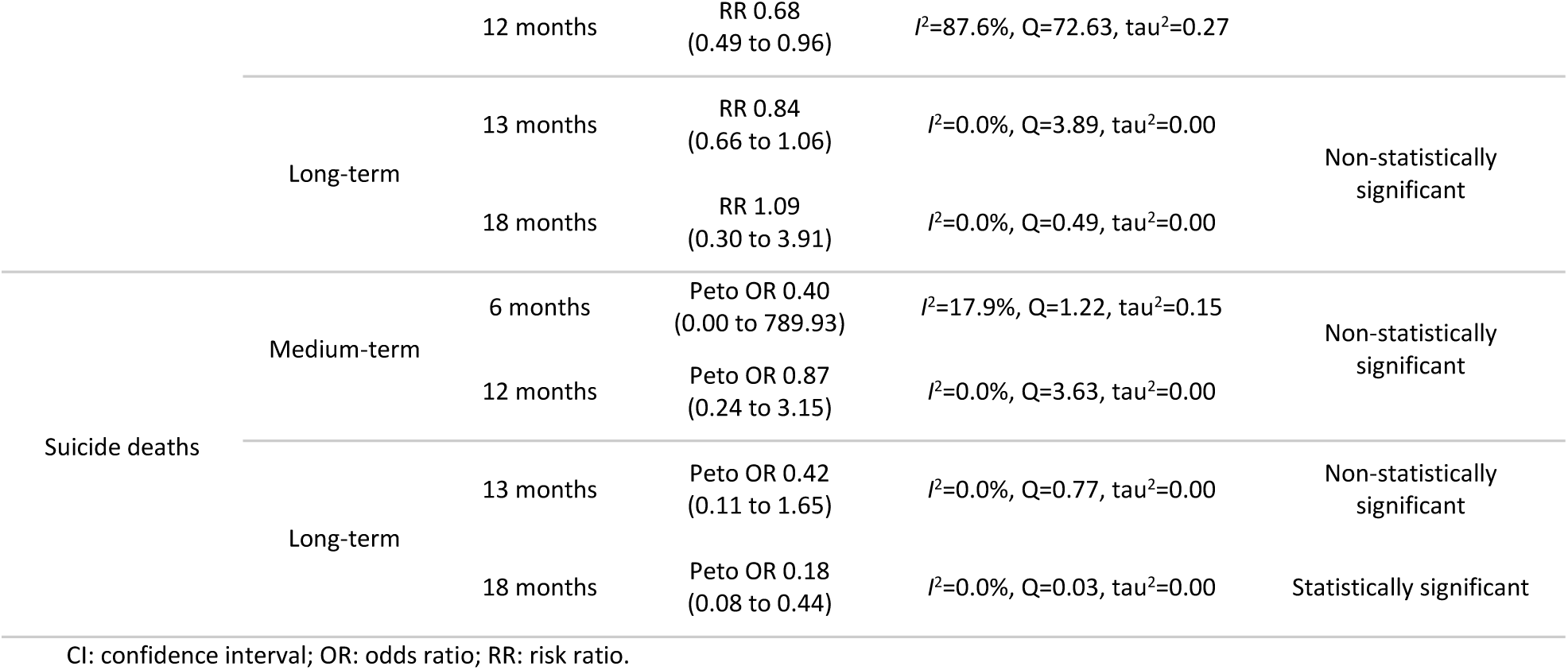
Summary of intervention effects grouped by follow-up period (short-, medium-, and long-term).

### Results of narrative synthesis

Effects of Telephone Contact Interventions on Suicidal Behaviours A total of 15 articles examined the effectiveness of telephone follow-up interventions in the prevention of suicidal ideation, suicide attempts, and suicide deaths [59–61, 64, 65, 67, 68, 70, 71, 75, 76, 78, 80, 81, 83]. The studies by Cebrià *et al*. [59], Exbrayat *et al*. [65], Malakouti *et al*. [71], Plancke *et al*. [78], and Vaiva *et al*. [80] reported differences in suicide reattempts in the intervention group compared to the control group, and a significant effect of the intervention group in reducing the rate of reattempts. Cebrià *et al*. [60], after a 60-month follow-up, indicated that the significant differences found at 12 months were not maintained in either group in terms of suicide reattempts (31.4% vs. 34.4%, *p*=0.40).

Demesmaeker *et al.* [64] examined the impact of the Vigilans programme by comparing interventions of a telephone call only between the 10th and 21st days after discharge, postcards at months 2–5 (if unreachable), a combination of both (if in suicidal crisis), and TAU. The Kaplan-Meier method, illustrating the time to the first suicide reattempt within 6 months, showed the highest survival probability for individuals receiving only a telephone contact and the lowest for those receiving TAU.

Cedereke *et al*. [61] and Mousavi *et al*. [76] found significant reductions in suicidal ideation, regardless of the intervention condition. On the other hand, Wei *et al*. [83] compared a telephone intervention with a cognitive therapy group and a control group, showing reductions in the number of suicide reattempts between the intervention and control groups (1.3% vs. 6.5%, *p*=0.08), although the differences were marginally significant. No statistically significant differences in suicidal ideation were shown between the three groups at any assessment point (baseline: *p*=0.43, 3-month: *p*=0.72, 6-month: *p*=0.75, 12-month: *p*=0.67).

Gabilondo *et al*. [67], Goñi-Sarriés *et al*. [68], López-Goñi & Goñi-Sarriés [70], Mousavi *et al*. [75], Mousavi *et al*. [76], and Vaiva *et al*. [81] reported no significant differences between intervention and control groups for suicide reattempts. In addition, Goñi-Sarriés *et al*. [68] informed that there were no significant differences in suicide deaths. More detailed information is in Table 1.

#### Effects of Text Message Interventions on Suicidal Behaviours

Comtois *et al*. [63] conducted a follow-up text-messaging intervention with soldiers and marines considered at risk of suicide. Eleven contacts were made by mental health clinicians, with participants’ text message responses monitored 24 hours a day. The authors reported no significant effect of the intervention on the presence or severity of current suicidal ideation; furthermore, the intervention did not significantly decrease the odds of a suicide risk incident during the 12-month follow-up period.

#### Effects of Multi-component Interventions on Suicidal Behaviours

A total of 12 articles examined the effectiveness of a multi-component synchronous telepsychiatry strategies– e.g., 1-hour psychoeducational session plus telephone contacts– in the prevention of suicidal ideation, suicide attempts, and suicide deaths [56–58, 62, 66, 69, 72–74, 77, 79, 82]. Miller *et al*. [73] evaluated the ED-SAFE intervention, which consisted of a risk screening and seven post-ED telephone calls. No significant differences in risk reduction were found between the TAU and screening phases (22.9% vs. 21.5%, *p*=0.99). Mouaffak *et al*. [74] delivered postcards, in combination with the possibility of calling a 24-hour telephone, and with telephone follow-up at 2 weeks, 1, and 3 months after discharge from the ED. No significant differences were found between the intervention and control groups for suicide reattempts (14.5% vs. 14%, *p*=0.89). Messiah *et al*. [72] evaluated the effectiveness of the AlgoS Algorithm, which consisted of a telephone follow-up at 10 and 21 days, for multi-attempters, or a crisis card, for first-attempts, versus TAU. There was no significant difference for multi-attempters in the intervention group compared to the control group for suicide reattempts at 6 (RR 0.84, 95% CI 0.57 to 1.25) and 14 months (RR 1.00, 95% CI 0.73 to 1.37).

Burback *et al.* [58] investigated the impact of 12 web-based, therapist-administered EMDR sessions targeting experiences associated with suicidal ideation. Both EMDR and TAU group participants reported significant differences (*p*=0.009) in suicide ideation scores between baseline and 4-month follow-up (EMDR group baseline: *M*=20, *SD*=7.6; EMDR group 4-month: *M*=13, *SD*=9.6; TAU group baseline: *M*=20, *SD*=7.6; TAU group 4-month: *M*=13, *SD*=9.6). Additionally, EMDR group participants reported fewer serious adverse events compared to TAU group participants.

Amadéo *et al*. [56], Bertolote *et al*. [57], Fleischmann *et al*. [66], and Hassanzadeh *et al*.[69] investigated the efficacy of BIC using a brief 1-hour educational intervention combined with telephone contacts at 1, 2, 4, 7, 11 weeks, 4, 6, 12, and 18 months delivered by physicians, psychiatric nurses, psychologists, psychiatrists, or social workers. No significant reduction was found in the intervention group compared to the control group for suicide reattempts, although, a significant reduction in suicide deaths in the intervention group compared with the control group (0.2% vs. 2.2%, *p*<0.001) at 18 months was reported in the study by Fleischmann *et al*. [66]. In a similar intervention protocol, Riblet *et al*. [79] contacted participants by telephone or video over 3 months, mailed educational materials, and performed a 1-hour educational intervention on suicide prevention also by telephone or video. A reduction in suicidal ideation was found in the intervention group compared with the control group at 1 and 3 months.

Christensen *et al*. [62] conducted a web-based CBT intervention combined with telephone follow-up compared with a CBT-only intervention and a TAU group. CBT was delivered in five modules, and telephone contacts were provided weekly by a 24-hour telephone counselling service. Significant reductions in suicidal ideation, in the telephone call condition, at 1-month post-intervention (*p*=0.003), 6-month follow-up (*p*=0.03), and 12-month follow-up (*p*=0.01) were found. Similarly, Ward-Ciesielski *et al.* [82] conducted a single-session Dialectical Behaviour Therapy Skills-Based Intervention (DBT-BSI) and telephone follow-up at 1, 4, and 12 weeks, compared with a control group. A significant reduction in levels of suicidal ideation was reported in both conditions, although there were no significant differences between conditions for suicidal ideation and the number of suicide attempts. O’Connor *et al*. [77] evaluated the effects of the SAFETEL intervention against TAU. A safety planning intervention (SPI) session and five weekly optional telephone calls were conducted. No significant differences were found between the intervention group and the control group for suicide attempts (Incidence Rate Ratio; IRR 1.10, 95% CI 0.43 to 2.79, *p*=0.85).

#### Effects of Synchronous Remote-Based Interventions on Treatment Adherence

Nineteen studies reported on outcomes relating to treatment attendance [56–61, 64, 66–72, 77, 80–83]. Concerning the moderating effect of adherence on the effectiveness of the intervention: Gabilondo *et al*. [67] observed that the intervention group showed significantly higher adherence to outpatient follow-up (*p*<0.001); in addition, Hassanzadeh *et al*. [69] revealed an increase in patients’ perceived need for support (*p*<0.001) and positively influenced attitudes towards seeking support from services and family. Moreover, Vaiva *et al*. [80] reported that participants in the intervention group discussed their suicide attempts with their primary care physician more frequently than controls.

According to Cebrià *et al*. [60], suicide attempters tend to be ambivalent to treatment, do not attend treatment, or drop out prematurely; complementarily, López-Goñi & Goñi-Sarriés [70] showed in their study that completion of the Telephone Follow-up Programme (TFP) correlated with a reduction in suicidal behaviours. Along the same lines, Demesmaeker *et al.* [64] highlight the importance of addressing the specific challenges faced by people who are unwilling to enter treatment in the context of suicide prevention interventions. Interventions may not uniformly benefit all patients, possibly due to differences in baseline suicidality or programme adherence.

Ward-Ciesielski *et al*. [82], and Wei *et al*. [83] also mention that the 75% retention rate underscores the acceptability and effectiveness of the study in engaging a historically hard-to-reach and difficult-to-retain population. As mentioned by Messiah *et al*. [72] telephone calls could complement a multimodal strategy, providing a surveillance tool to drive more aggressive interventions. Nevertheless, technical challenges, lack of access to computers, or lack of privacy could prevent the enrollment of some individuals, as noted by Burback *et al.* [58].

Fleischmann *et al*. [66], Gabilondo *et al*. [67], and Demesmaeker *et al.* [64] mention that BIC operates similarly to psychosocial counselling, providing temporary social support and fostering feelings of connectedness through systematic follow-up contacts. BCI may help to reduce social isolation and facilitate the progressive engagement of patients who may initially be reluctant to participate in structured psychotherapies.

Concerning the predictor or moderator variables of adherence: Amadéo *et al*. [56], López-Goñi & Goñi-Sarriés [70], O’Connor *et al*. [77], Vaiva *et al*. [80], and Vaiva *et al*. [81] concurred that patients lost to follow-up were more likely to be male. Gabilondo *et al*. [67], Goñi-Sarriés *et al*. [68], López-Goñi & Goñi-Sarriés [70], Vaiva *et al*. [80], and Vaiva *et al*. [81] concluded that the intervention could be more effective for higher-risk populations requiring additional support; in these studies, patients who completed PTF demonstrated a higher severity of suicide attempts, greater psychopathological severity, used more violent methods, and experienced predominantly affective disorders. In contrast, Ward-Ciesielski *et al*. [82] found that participants lost to follow-up reported higher levels of depression and anxiety. In addition, Demesmaeker *et al.* [64] reported that post-traumatic stress disorder and generalised anxiety disorder appear to be predictors of non-adherence to the programme. On the other hand, four studies [66, 71, 77, 83] discussed stigma around suicide as a factor that may affect patients’ attitudes towards seeking support from mental health services and family. More detailed information is in Supplemental File 6: Table S4.

#### Causes of heterogeneity among study results

The investigations on the possible causes of heterogeneity between the outcomes of suicidal ideation (*I*^2^=59.4% at 1 month, *I*^2^=69.3% at 6 months, *I*^2^=51.9% at 12 months) and suicide attempts (*I*^2^=85.4% at 6 months, *I*^2^=87.6% at 12 months) of the selected studies raise the pertinence of qualitatively synthesising the results due to heterogeneity in: a) the evaluation measures time frame of the outcomes (see Supplemental File 7. Table S5), b) the protocol of intervention design and the characteristics of the intervention and control group– i.e., target effect, measures of effect, method of implementation, administrator-participant interaction, periodicity, number and duration of the intervention, professional delivering the intervention, setting, participants, etc.– (see Supplemental File 5. Table S3), and c) assessment of RoB and study design.

### Subgroup analysis

Subgroup analysis was conducted based on length of intervention, type of treatment, research design, and RoB assessment, if feasible [95]. Analysis revealed, for 1-month suicidal ideation outcomes, that adjustment for confounding variables altered the direction of the effect, with adjusted analysis indicating a negative effect (Hedges’ *g* -0.41, 95% CI -1.12 to 0.31) and unadjusted analysis showing a small positive effect (Hedges’ *g* 0.17, 95% CI 0.00 to 0.33). On the other hand, the results suggest that telephone contact interventions tend to show greater effectiveness in reducing suicide attempts at the 6-month follow-up (RR 0.37, 95% CI 0.22 to 0.62) compared to multi-component interventions (RR 0.99, 95% CI 0.59 to 1.65). Additionally, nRCTs demonstrated a stronger effect (RR 0.31, 95% CI 0.18 to 0.53) compared to RCTs (RR 0.84, 95% CI 0.60 to 1.18) at 6 months follow-up. More detailed information is in Supplemental File 8: Table S6, S7 and S8.

### Publication bias

Publication bias was examined using both funnel plots and Egger’s test. The visual inspection of the funnel plots suggested no evidence of publication bias (see Supplemental File 9: Figure S1, S2, and S3); supported by the results of Egger’s test, which indicated absence of evidence for publication bias in the included primary studies for suicidal ideation (*p*=0.92), suicide attempts (*p*=0.24), and suicide deaths (*p*=0.12).

### Certainty of evidence

The overall certainty of the evidence, according to the Grading of Recommendations Assessment, Development, and Evaluation (GRADE) [53], was moderate for RCTs reporting suicidal ideation and suicide death, and low for RCTs and nRCTs reporting suicide attempts. The main reasons for downgrading were risk of bias, inconsistency, and imprecision in the results (Supplemental File 10. Table S9).

## DISCUSSION

The aim of the study was to assess the effectiveness of synchronous remote interventions in relation to three indicators related to suicidal behaviour: suicidal ideation, suicide attempt, and death by suicide. The results of our systematic review and meta-analyses are largely consistent with previous findings on the effectiveness of telematic interventions in preventing suicidal behaviour. In line with the Shoib *et al*. study [36], our conclusions reinforce the value of telepsychiatry in reducing suicide reattempt rates, although with heterogeneity in effects. Nevertheless, our research represents a significant advancement, offering more recent findings, a greater number of studies, additional variables, a wider range of telepsychiatry interventions, and an analysis of the temporal evolution of the intervention effects, providing a more nuanced understanding of their impact. Furthermore, we specifically analysed suicidal ideation as a possible mediator of the effects of these interventions, providing new insights into the mechanisms that might explain their effectiveness.

### Meta-analyses: Effectiveness of synchronous remote-based interventions

The evidence suggests that, although synchronous remote-based interventions appear to reduce suicide attempts and, to some extent, deaths by suicide, their impact on suicidal ideation remains limited. The results seem to indicate that interventions have a clear effect in reducing suicide attempts, with an inverted U-shaped evolution over time, showing a peak of effectiveness at 6 months (-44%, 95% CI -66% to -5% in the intervention group compared to the control group) which subsequently declines. On the contrary, the results of the study do not show statistically significant effects on suicidal ideation, which exhibit a small effect size throughout the four evaluation moments. From the set of studies analysed in relation to deaths by suicide, it is observed that the confidence intervals cover very large ranges of odds ratios.

This is probably because they refer to a very low-frequency phenomenon, which exceeds the sampling power of conventional empirical clinical trial studies. The juxtaposition of several of these studies in a meta-analysis also does not solve the problem of lack of sampling power for this rare occurrence. Likewise, the difficulty of obtaining reliable information on suicide deaths is highlighted, which adds another challenge to reaching consistent conclusions. Moreover, in some of our meta-analyses (e.g., 6-month time point in Figure 5), the use of the Knapp-Hartung adjustment in random-effects models led to unusually wide confidence intervals when a few studies were included (i.e., *n*=2). In situations like the one highlighted above, a high level of uncertainty is generated, and the results clearly do not reflect the current weight of the evidence in an intuitive sense.

In addition to the marked differences in the effect sizes of synchronous remote-based interventions on the different indicators, their evolution over time differs, which can be interpreted as a probable consequence of variations in the studies used for the evaluation of the three indicators across the time points, compromising comparability. But it could also be revealing differential effects of the interventions on each of the outcomes. In particular, the marked difference in the effect sizes observed between suicidal behaviour and suicidal ideation, which appears to be consistent across the assessment moments, along with the differing evolution of these outcomes over time, suggests that the clear behavioural effect of the intervention is not cognitively mediated. Most theoretical models of suicidal behaviour [96] assume that it is constituted by a continuum of processes ranging from passive ideation and planning to death through suicide attempt. Considering this sequencing, the apparent effect of synchronous remote-based interventions on suicide attempts— but not on suicidal ideation— suggests the probable involvement of an inhibitory mechanism. This mechanism may act directly on behaviour, temporarily counteracting the influence of suicidal ideation on suicidal actions, while the ideation itself probably persists as a latent risk factor. In this sense, it should be considered that a key aspect of synchronous remote-based interventions is that they provide the person at risk of death by suicide with regular, high-quality social interaction and support, which is related to the concept of ‘connectivity’ discussed further. The discrepancy may also be explained by the nature of the interventions, which often emphasise external support, monitoring, and engagement rather than targeting the cognitive and emotional processes underlying suicidal thoughts. While such strategies may enhance safety and prevent behavioural outcomes like suicide attempts, they may not sufficiently address internal experiences. A recent meta-analysis of safety planning-type interventions for suicide prevention [97] also finds differences in reducing suicidal attempts but not suicidal ideation. The mechanism behind the effectiveness of these interventions lies in risk management, preventing the transition from suicidal ideation to suicidal action, even though no significant reduction in suicidal thoughts was observed.

Despite differences in the trends over time of the three indicators evaluated, some commonality exists since they generally tend to improve over time, probably reflecting a cumulative effect of the intervention that plateaus around 12-13 months. However, this finding should be taken with caution as the studies included in the different evaluation moments are not always the same, which may partially explain apparent changes in effect size over time.

### Quality, comparability, and integrability of the studies included in the meta-analyses

There are various aspects of the nature of the studies analysed, as well as the methodology followed in the meta-analyses, that condition the interpretation of the results. Firstly, of the 28 studies analysed, only eight (29%) had a low RoB, including seven randomised controlled trials (37%) and one non-randomised trial (11%). Additionally, unexplained statistical heterogeneity across studies warrants further investigation to determine effects.

Secondly, although the selected studies share key commonalities– namely, all being clinical trials and involving synchronous, remote-based interventions– there is substantial variability in their action protocols and evaluation points. Significant variations were found in the procedure of the intervention, and comparison group– i.e., periodicity, number of contacts, moment of assessment, content, practitioner, setting, type of usual treatment, etc.–, as well as in the methodology– i.e., study design and RoB– of the reviewed studies. Heterogeneity of results could be due to differences in the length of follow-up, the methods used in the different programmes, variability in the number of sessions, or different patient inclusion criteria. This heterogeneity complicates the integration of findings into a meta-analysis. However, it has been partially addressed by conducting specific meta-analyses based on temporal groupings, which enhance comparability while also assessing the dynamic evolution of the intervention’s therapeutic effects. Replication of these interventions according to a standardised protocol would facilitate more consistent results [98].

### Narrative synthesis

Firstly, the neologism ‘connectivity’ seems to serve as a central concept in the majority of studies [35]. In addition, the importance of a high frequency of follow-up contacts, earlier intervention, and involvement in the most critical period– i.e., 6 months post-discharged from ED– is also underlined [10, 13, 35, 80]. Reducing loss to follow-up is a critical challenge in suicide prevention, especially during the post-discharge period [13, 99], given historical problems with treatment completion among people involved in suicidal behaviour [100–103]. According to Gabilondo *et al*. [67] BIC reduces loneliness and improves adherence to routine treatment. Along the same lines, Ward-Ciesielski *et al*. [82] reported that a notable percentage of participants engaged with mental health services, or initiated psychotherapy, or pharmacotherapy during the synchronous remote intervention follow-up, suggesting a significant benefit of these interventions for a treatment-unengaged population.

### Recommendations, implications for clinical practice, and future research

Following the synthesis of information and the articles collected on the topic of the review, the authors would like to provide a series of recommendations to standardise the implementation and development of these interventions. Firstly, the authors of future studies should consider describing in detail a minimum set of component characteristics of the intervention and comparison groups– particularly for the TAU group– as there is considerable heterogeneity between studies. A comprehensive quality checklist is presented in Supplemental File 11. Table S10.

Secondly, as concluded in the systematic review by Noh *et al*. [33] more structured and theory-based interventions for the prevention of suicidal behaviours should be conducted and examined, emphasising the importance of a conceptual and theoretical framework in the development of interventions, and increasing measurable impacts on key outcomes in complex therapies [104]. A universal intervention design protocol should be adopted according to the characteristics of the studies in which an effect has been observed and the recommendations of the authors of previous studies on the topic [59, 62, 65, 66, 71, 78, 80, 81]. In summary, the preferred approach is a RCT design that delivers early interventions within a maximum of 7 days post-ED discharge with long-term follow-up of approximately 12 months, often as concomitant therapy to TAU– i.e., emergency medical care and follow-up based on the severity of suicidal behaviour; psychopharmacology, or psychotherapy– conducted by a mental health professional to enhance suicide risk assessments, clinician-patient connection, and adherence to routine treatment.

Overall, the findings emphasise the importance of standardising interventions to enhance both research comparability and clinical application. Moreover, the results open up several avenues for further investigation into effectiveness, underscoring the need to standardise the outcomes to be assessed, the timing of evaluations, and the measurement instruments. Addressing the impact of this intervention on suicide deaths will require much larger cohorts, which in turn implies improving the protocols for recording this indicator to ensure data reliability. Finally, the suspicion that synchronous remote-based interventions could have an effectiveness on the reduction of suicide attempts not mediated by a reduction in suicidal ideation makes it necessary to focus the future of research on the processes that mediate the effectiveness of interventions, which may be crucial for improving their efficacy and cost-effectiveness. In particular, future research could benefit from integrating components designed to promote cognitive change and emotional regulation in order to more effectively influence ideation.

### Strengths and limitations

The primary limitation is the significant heterogeneity observed in the overall meta-analyses and subgroup analyses, which may be attributed to the small number of studies included in some analyses or the substantial variability between their results. In addition, using ED data records or the authorities’ forensic register may lead to the possibility of under- reporting the actual number of non-fatal suicidal behaviours [105]. Furthermore, suicide is a rare event, making the design of studies with high statistical power particularly difficult. Finally, the included trials used intervention and comparison groups that received TAU; these treatments are often different across studies. Relatedly, although study characteristics were described, the lack of standardised reporting across research limited further stratification.

The comprehensive literature review and the identification of key characteristics for improving future study designs are fundamental strengths. Other strengths of the study are the number of databases explored, the reporting according to PRISMA guidelines, the validation of the boolean searches according to PRESS guidelines, and the inter-reviewer consensus in the different phases of screening, data extraction, and RoB assessment.

## CONCLUSIONS

Innovation in the prevention and management of suicidal behaviour is a therapeutic challenge [106]. The current research outlines emerging evidence on suicide prevention through telemedicine in reducing suicidal behaviours. The main conclusion of the study is that the results seem to indicate that synchronous remote-based interventions demonstrate a potential effect in reducing suicide attempts and suicide deaths, although their influence on suicidal ideation appears limited. The mechanisms underlying these effects remain unclear but may be related to the promotion of social bonding or ‘connectedness’. The difficulties in studying the effectiveness of such preventive interventions are reflected by the great heterogeneity of studies to date and the problems in obtaining accurate information on suicidal behaviours in the absence of reliable registries [107]. Finally, an indirect positive effect on the treatment of patients at risk of death by suicide seems to derive from the increase in adherence fostered by remote synchronous interventions.

## Data Availability

Data are available in a public, open access repository. The dataset is available in CORA RDR, https://doi.org/10.34810/data1310

## ETHICS AND DISSEMINATION

Ethics approval is not needed, as systematic reviews are based on published studies. The results will be disseminated through peer-reviewed publications.

## ETHICS STATEMENTS

Patient consent for publication Not applicable.

## AUTHOR CONTRIBUTIONS

AS is the guarantor. LC, JML, DP, AC, and AS: Writing - Original Draft. LC, AS, MPJ, JPS, and CM: Software. LC, JML, DP and AS: Project administration, Supervision. All authors: Conceptualization, Methodology, Writing - Review & Editing. JML, AS, JPS, and CM provided statistical expertise. DP and AC provided expertise on suicidal behaviours. All authors approved the final manuscript.

## AI USE

Google Translate was used to check (and modify where necessary) the linguistic quality and suitability of the manuscript. No other AI tools were used in either the study or the writing of the manuscript.

## ACKNOWLEDGEMENTS

Authors’ thanks to Guillem Cebrián, director of the Library at Parc Taulí University Hospital and head of the Unitat de Gestió del Coneixement de l’Institut d’Investigació i Innovació Parc Taulí (I3PT-CERCA), for his support in the refinement of the search strategies. DP thanks the support of Spanish Ministry of Science and Innovation/ISCIII/MICIU/FEDER (PI21/01148); the Secretaria d’Universitats i Recerca del Departament d’Economia i Coneixement of the Generalitat de Catalunya (e-MH-PEMN – 2021 SGR 01431); the CERCA programme / Generalitat de Catalunya; the Instituto de Salud Carlos III; and the CIBER of Mental Health (CIBERSAM CB 19/09/00029). The research has been previously presented at a conference and has been published as a conference abstract [108].

## FUNDING

This research was funded by the Instituto de Salud Carlos III, Subdirección General de Evaluación y Fomento de la Investigación (ISCIII), Ministerio de Ciencia, Innovación y Universidades (MICIU) co-funded by the European Union, Fondo Europeo de Desarrollo Regional (FEDER) (grant number PI21/01148 — COGNISUI) and MCIN/AEI/10.13039/501100011033 ’ERDF A way of making Europe’ (grant number PID2022-141403NB-I00). Fundació Institut d’Investigació i Innovació Parc Taulí (I3PT-CERCA), SENDO PI21/01148 (Fondo de Investigación Sanitaria-ISCIII/ FEDER), Instituto de Salud Carlos III. The funders had no role in the design of the study; in the collection, analysis, or interpretation of data; in the writing of the manuscript; or in the decision to publish the results. The Department of Mental Health at the Parc Taulí University Hospital and the Institut d’Investigació i Innovació Parc Taulí (I3PT-CERCA) are the sponsors.

## COMPETING INTERESTS

DP has received grants and served as a consultant or advisor for Rovi, Johnson & Johnson, and Lundbeck. The other authors declare no conflicts of interest.

## PATIENT AND PUBLIC INVOLVEMENT

Patients and/or the public were not involved in the design, conduct, reporting, or dissemination plans of this research.

## PATIENT CONSENT FOR PUBLICATION

Not applicable.

## PROVENANCE AND PEER REVIEW

Not commissioned; externally peer reviewed.

## DATA AVAILABILITY STATEMENT

Data are available in a public, open access repository. The dataset is available in [Dataset] Comendador L; Jiménez-Villamizar MP; Sanabria-Mazo, JP, *et al*. Data from: Effect of synchronous remote-based interventions on suicidal behaviours: systematic review and meta-analysis. CORA Repository, February 9, 2025. https://doi.org/10.34810/data1310 [109]

## SUPPLEMENTAL MATERIAL

Supplemental File 1. PRISMA 2020 Checklist (DOCX 32 KB).

Supplemental File 2. Amendments made to the original protocol (DOCX 11 KB).

Supplementary File 3. Search strategy for databases, registers, and websites (DOCX 13 KB). Supplemental File 4. List of articles excluded after full-text screening (DOCX 23 KB).

Supplemental File 5. Information on the components and characteristics of the intervention and comparison groups (DOCX 20 KB).

Supplemental File 6. Information on therapeutic adherence (DOCX 15 KB).

Supplemental File 7. Evaluation measures the time frame of the selected studies (DOCX 14 KB). Supplemental File 8. Subgroup analysis (DOCX 27 KB).

Supplemental File 9. Publication bias (DOCX 130 KB). Supplemental File 10. GRADE (DOCX 689 KB).

Supplemental File 11. Report of the minimum set of component characteristics (DOCX 11 KB).

## OPEN ACCESS

This is an open-access article distributed by the Creative Commons Attribution Non-Commercial (CC BY-NC 4.0) license, which permits others to distribute, remix, adapt, build upon this work non-commercially, and license their derivative works on different terms, provided the original work is properly cited, appropriate credit is given, any changes are made indicated, and the use is non-commercial. See: http://creativecommons.org/licenses/by-nc/4.0/.

## ORCID iDs

Laura Comendador Vázquez https://orcid.org/0000-0002-5221-4794 María P Jiménez-Villamizar https://orcid.org/0000-0003-2264-7422 Josep-Maria Losilla https://orcid.org/0000-0002-5140-5847

Juan P Sanabria-Mazo https://orcid.org/0000-0003-1688-435X Corel Mateo-Canedo https://orcid.org/0000-0002-0620-9257 Antoni Sanz Ruíz https://orcid.org/0000-0002-7952-4477

Ana-Isabel Cebrià Meca https://orcid.org/0000-0002-2632-8130 Diego Palao Vidal https://orcid.org/0000-0002-3323-6568

## Author’s Note

Laura Comendador Vázquez, MSc, PhD student. ORCID 0000-0002-5221-4794

María P. Jiménez-Villamizar, MSc, PhD student. ORCID 0000-0003-2264-7422

Josep-Maria Losilla, PhD. ORCID 0000-0002-5140-5847

Juan P. Sanabria-Mazo, PhD. ORCID 0000-0003-1688-435X

Corel Mateo-Canedo, PhD. ORCID 0000-0002-0620-9257

Antoni Sanz Ruíz, PhD. ORCID 0000-0002-7952-4477

Ana Isabel Cebrià Meca, PhD. ORCID 0000-0002-2632-8130

Diego J. Palao Vidal, MD, PhD. ORCID 0000-0002-3323-6568

## Notes

### Competing Interest Statement

D.P. has received grants and also served as a consultant or advisor for Rovi, Angelini, Janssen, Lundbeck and Servier. The other authors declare no conflicts of interest.

### Clinical Protocols

https://doi.org/10.1136/bmjopen-2023-075116

### Funding Statement

This research was funded by the Instituto de Salud Carlos III, Subdireccion General de Evaluacion y Fomento de la Investigacion (ISCIII), Ministerio de Ciencia, Innovacion y Universidades (MICIU) cofunded by the European Union, Fondo Europeo de Desarrollo Regional (FEDER) (grant number PI21/01148, COGNISUI) and MCIN/AEI/10.13039/501100011033 ERDF A way of making Europe (grant number PID2022-141403NB-I00). Fundacio Institut d'Investigacio i Innovacio Parc Tauli (I3PT-CERCA), SENDO PI21/01148 (Fondo de Investigacion Sanitaria-ISCIII/ FEDER), Instituto de Salud Carlos III. The funders had no role in the design of the study; in the collection, analysis, or interpretation of data; in the writing of the manuscript; or in the decision to publish the results. The Department of Mental Health at the Parc Tauli University Hospital and the Institut d'Investigacio i Innovacio Parc Tauli (I3PT-CERCA) are the sponsors.

### Summary of Updates

This the final preprint version accepted to be published in the journal BMJ Open. Compared with the first (current) version uploaded to MedRXiV, it includes the ammendments done in the revision process of this journal.

